# A systematic review of low-dose interleukin-2 for the treatment of systemic lupus erythematosus

**DOI:** 10.1101/2022.12.02.22283038

**Authors:** Lara Bader, Onur Boyman, Miro E. Raeber

## Abstract

**Background:** Low-dose interleukin-2 (IL-2) shows promise as a treatment for restoring functional and numerical deficits of regulatory T (Treg) cells in patients with various autoimmune diseases. Several clinical trials testing low-dose IL-2 in systemic lupus erythematosus (SLE) have been completed, with a comprehensive review of these trials currently lacking.

**Objective:** We aimed to conduct a systematic synthesis of findings from clinical trials regarding the clinical efficacy and safety of using low-dose IL-2 in patients with SLE. Furthermore, we intended to determine the sensitivity of different responder indices for IL-2-induced clinical improvement in SLE.

**Methods:** Following the Preferred Reporting Items for Systematic Reviews and Meta-Analyses, we searched the Scopus and MEDLINE databases for articles reporting trials testing low-dose IL-2 in patients with SLE published between January 2010 and September 2022. To evaluate the risk of bias, we applied a modified version of the Downs and Black assessment tool.

**Results:** We retrieved 1,018 articles, six of which we analyzed including four open-label studies and two randomized controlled trials following a detailed review process. The studies included a total of 230 patients, of which 150 received low dose IL-2 and 80 received placebo. Although the open-label studies demonstrated an expansion of Treg cells that coincided with a clinical response, the predefined primary endpoints for clinical efficacy were not achieved in the randomized controlled trials. In general, treatment with low-dose IL-2 appears to be safe and tolerated well.

**Conclusion:** Low-dose IL-2 therapy appears to be a promising strategy for treating SLE; however, larger trials are still necessary to assess clinical responses in patients with such a heterogeneous disease.

## I. Introduction

Systemic lupus erythematosus (SLE) is an autoimmune disease that significantly impacts the morbidity and mortality of affected patients with an estimated 10-year survival rate of 70%. SLE disproportionally affects young women of childbearing age and has high geographic variability in its prevalence that ranges from 20 to 150 per every 100,000 people (1). Characterized by autoantibodies directed against nuclear self-antigens, SLE was initially considered to be a prototypical disease mediated by B cells; however, progress in characterizing the disease has shifted that perspective (2). However, recent success of chimeric antigen receptor T cells application in 8 SLE patients has refueled the B cell-centric view (3). Although recent work characterizing SLE in large cohorts following multiomics approaches has provided unprecedented insights into this multifactorial disease (4, 5), its complex pathogenesis remains poorly understood. SLE is associated with heterogeneous manifestations affecting different organ systems, including the musculoskeletal system, the central nervous system and the skin, lungs, and kidneys (6). Current treatment approaches remain primarily based on broad immunosuppression (7), and in the past 60 years, only the anti-B cell activating factor antibody Belimumab and the anti-type I interferon receptor antibody Anifrolumab have been approved by the U.S. Food and Drug Administration (FDA) for treating SLE (8–10). Meanwhile, many other attempts to use biologics that succeeded in other indications have failed for the treatment of SLE.

Regulatory T (Treg) cells are indispensable for maintaining peripheral tolerance, as the development of severe autoimmunity in Treg-deficient mice and patients has shown (11). Research on Treg cells revealing a functional and/or numerical deficit in different autoimmune diseases (12) has motivated initial clinical trials testing the use of low-dose interleukin-2 (IL-2) to expand Treg cells for the treatment of cryoglobulinemic vasculitis (13), chronic graft-versus-host disease (14), and other indications (15). The underlying mechanism can be explained by two different primary configurations of the IL-2 receptor. On the one hand, a dimeric receptor consisting of the IL-2 receptor β (CD122) and the common γ chain (CD132) is sufficient for signal transduction and, due to its intermediate affinity for IL-2, is stimulated by high doses of IL-2. On the other hand, the trimeric IL-2 receptor additionally including the IL-2 receptor α (CD25), with a 10- to 100-fold higher affinity for IL-2, is activated by already low doses of IL-2 (16). Because the dimeric IL-2 receptor is primarily expressed on memory-phenotype effector T cells and the trimeric receptor primarily on Treg cells, the exogenous application of low-dose IL-2 preferentially stimulates Treg cells.

This has spurred interest in investigating Treg cells in SLE, research on which has revealed a functional defect of Treg cells also present in SLE and has led to the first trials testing low-dose IL-2 in this disease (17). To date, several open-label studies and randomized controlled trials (RCTs) have examined the efficacy and safety of using low-dose IL-2 in patients with SLE. Therein, one challenge has been the difficulty of accurately measuring the disease’s severity and responses to treatment due to SLE’s complex presentation and heterogeneity. Several clinical scores have been used to measure clinical response, including the SLE Disease Activity Index (SLEDAI), the British Isles Lupus Assessment Group (BILAG) index, the Physician Global Assessment (PGA) score, and composite outcome scores including 4-point improvement on the SLE Responder Index (SRI-4) and the BILAG-Based Composite Lupus Assessment (BICLA) (18–22). The composite endpoints on the SRI-4 and BILAG are particularly relevant in developing drugs for SLE, as they have been accepted by the U.S. FDA for the approval of belimumab and anifrolumab, respectively (9, 10). With the SRI-4 based on the SLEDAI, including serological markers, and the BICLA based on the BILAG, which is considered to be relatively sensitive to clinical improvements, a lingering question for studies testing low-dose IL-2 or even improved IL-2 formulations in SLE concerns which endpoints to choose to complete a successful trial. Thus, in our systematic review, we aimed to summarize the clinical efficacy and safety of using low-dose IL-2 in patients with SLE and to determine the most sensitive index for IL-2-induced clinical improvement in SLE.

## II. Methods

### a. Study design and protocol registration

We registered our study’s protocol in the International Prospective Register of Systematic Reviews (Registration No. CRD42022328709) and followed the updated Preferred Reporting Items for Systematic Reviews and Meta-Analyses guidelines (PRISMA) (23).

### b. Search strategy

We searched the Scopus and MEDLINE databases for articles reporting clinical trials using IL-2 to treat SLE published in German or English between 01.01.2010 and 30.09.2022. The search terms were as follows:

MEDLINE: (‘Interleukin 2’ AND Lupus) AND ((“2010/01/01“[Date - Publication] : “2022/09/30“[Date - Publication]));

Scopus: (TITLE-ABS-KEY(interleukin-2) AND TITLE-ABS-KEY(lupus)) AND PUBYEAR > 2009 AND ( LIMIT-TO ( DOCTYPE,”ar” ) OR LIMIT-TO ( DOCTYPE,”le” ) ) AND ( LIMIT-TO ( LANGUAGE,”English” ) OR LIMIT-TO ( LANGUAGE,”GERMAN” ) ) AND (LIMIT-TO ( SRCTYPE,”j” ) ). We also examined a manuscript deposited on a preprint server, which was published in the journal *Immunity* in 2024 (24).

### c. Eligibility criteria

We included articles reporting open-label clinical trials or RCTs examining the effects of using low-dose IL-2 in patients diagnosed with SLE. We excluded articles reporting studies with nonhuman subjects, studies with non-standard treatment combinations (e.g., low-dose IL-2 combined with rapamycin), studies testing improved IL-2 formulations, studies focusing on specific disease manifestations (e.g., lupus nephritis), and studies that applied only a single cycle of low-dose IL-2 without clinical efficacy endpoints.

### d. Data extraction

Two authors (LB and MER) independently conducted the literature search and evaluated whether articles met the inclusion criteria. Duplicate search results were automatically eliminated using the literature management software Clarivate EndNote. Articles selected following title and abstract screening were retrieved for full-text review. For articles that met our inclusion criteria, we extracted the study’s characteristics (e.g., study design), the characteristics of patients (e.g., number of patients and demographic baseline data), and clinical results (i.e., clinical scores, biomarkers, and composite scores), which encompassed data from individual patients as well as metadata. The information gathered was cross-checked to ensure its accuracy and completeness. Discrepancies in eligibility-related decisions were discussed at each stage of the process until both authors reached consensus or by including a third author in the discussion.

### e. Risk of bias assessment

A modified Downs and Black checklist for assessment (25) was used to gauge the risk of bias (Table S2). The articles were scored by one author (LB) in four categories: reporting, external validity, internal validity, and power. This resulted in ranks of *high quality* (i.e., 23–28 points), *medium quality* (i.e., 15–22 points), and *low quality* (i.e., 0–14 points).

### f. Statistical analysis

To create graphs of the extracted data, including statistical analysis, we used GraphPad Prism version 9. Details about the statistical tests are provided in the text or figure legends when applicable.

### g. Principal summary measures and synthesis of results

Our systematic review aimed at providing a thorough summary of evidence about the efficacy and clinical safety of using low-dose IL-2 to treat SLE. To that end, we extracted clinical scores (i.e., SLEDAI, BILAG, PGA, SRI-4, and BICLA), information about IL-2-mediated Treg expansion, and safety data whenever available. Furthermore, our analysis was aimed to provide information on the sensitivity of the two composite endpoints SRI-4 and BICLA to measure IL-2-induced clinical response.

## III. Results

### a. Study selection

The process of selecting articles is illustrated in Fig. 1. In a preliminary search, we identified 1,018 articles, and after duplicates were removed, 877 articles remained for title and abstract screening. After initial screening, 22 articles were retrieved for full-text review, 17 of which did not meet our inclusion criteria. We added one article recently deposited on a preprint server, which was later published in the journal *Immunity* in 2024 (24). Thus, six studies were ultimately analyzed: two RCTs (26, 27) and four open-label studies (24, 28–30).

**Fig. 1.**
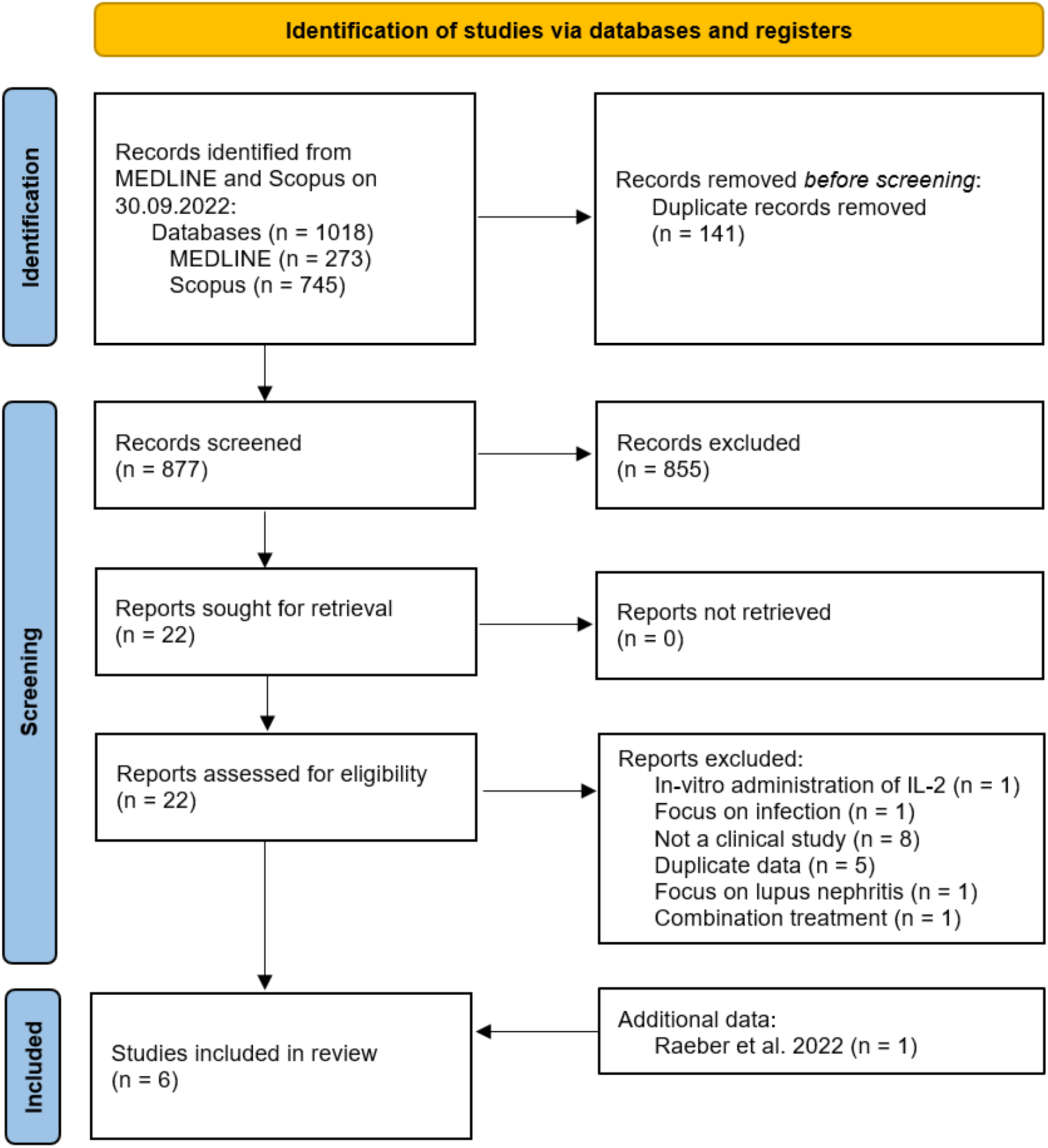
PRISMA 2020 flow diagram for systematic reviews.

In total, the reported trials included 230 patients, 150 of whom received low-dose IL-2, whereas 80 received a placebo. All patients were adults who had met the American College of Rheumatology’s diagnostic criteria for SLE updated in 1997 (31). Table 1 summarizes the baseline characteristics of the patients examined, Table 2 the primary results of the studies and Table 3 the reported adverse events. Our risk-of-bias assessment identified a higher risk of bias in the non-randomized studies (Table S2).

**Table 1:** Patient and study characteristics.

| Study | He 2016 (28) | He 2020 (34) |  | Humrich 2019 (30) | Humrich 2022 (26) |  |  |  | Rosenzweig 2019 (29) | Raeber 2024 (24) |
| --- | --- | --- | --- | --- | --- | --- | --- | --- | --- | --- |
| **Study Type** | Open-label uncontrolled | RCT |  | Open-label uncontrolled | RCT |  |  |  | Open-label uncontrolled | Open-label uncontrolled |
| **Time frame (until assessment of primary endpoint)** | 12 weeks | 12 weeks |  | 67 days | 12 weeks |  |  |  | 8 days (clinical endpoints 3 and 6 months) | 68 days |
| **Primary endpoint** | SRI-4 and safety | SRI-4 |  | Number of patients achieving a >100% increase in the proportion of CD25<sup>hi</sup>-expressing cells among CD3<sup>+</sup>CD4<sup>+</sup>FOXP3<sup>+</sup>CD127<sup>low</sup> Treg cells | SRI-4 |  |  |  | Change in relative concentration in peripheral blood Treg cells compared with baseline | Significant Treg cell expansion between baseline and day 68 after treatment initiation |
| **Primary endpoint met?** | Yes | No |  | Yes | Primary endpoint not met in ITT-population, met in PP-analysis |  |  |  | Yes | Yes |
| **Treatment details** | 3 cycles of subcutaneous IL-2, 1 MIU every other day for two weeks followed by a 2-week treatment break. Background treatment continued. | 3 cycles of 1 MIU IL-2 or placebo administered subcutaneously every other day for 2 weeks, followed by 2 weeks break. Background treatment continued. |  | 4 cycles of subcutaneous IL-2 for 5 consecutive days, followed by wash-out periods of 9, 16, and 16 days between cycles. Starting dose 1.5 MIU/day, increased to 3.0 or 4.5 if the expansion of Treg was insufficient or decreased. Background treatment continued. | 1.5 MIU of IL-2 or placebo administered subcutaneously every day for 5 days, then once every week from day 8 to week 12. Background treatment continued. |  |  |  | 1 MIU/day of IL-2 subcutaneously from day 1-5, then every 2 weeks from day 15-180. Background treatment continued. | 4 cycles of 1.5 MIU/day for 5 consecutive days, followed by a two-week treatment break. Background treatment continued. |
| **Treatment group** | Treatment | Treatment | Placebo | Treatment | Treatment ITT | Treatment PP | Placebo ITT | Placebo PP | Treatment | Treatment |
| **Number of patients** | 40 | 30 | 30 | 12 | 50 | 24 | 50 | 29 | 6 | 12 |
| **Female/Male** | 37/3 | 27/3 | 29/1 | 11/1 | 49/1 | 24/0 | 42/8 | 27/2 | 6/0 | 12/0 |
| **Age** | Median: 31<br>Range: 18-60 | Mean: 31.58<br>SD: 9.25 | Mean: 29.83<br>SD: 9.72 | Median: 38.5<br>Range: 26-61 | Mean: 41.7 | Mean: 41.3 | Mean: 40.4 | Mean: 38.2 | Mean: 46.3<br>Median: 45.0<br>Range: 27.0-67.0 | Median: 39.5<br>Range: 23-67<br>Mean: 42.2 |
| **Duration of disease** | Median: 5 years<br>Range: 0.5-20 years | Mean: 66.7 months<br>SD: 57.4 months | Mean: 63.6 months<br>SD: 59.9 months | Median: 9 years<br>Range: 2-35 years | Mean: 10.7 years<br>SD: 8.2 years | Mean: 13.7 years<br>SD: 9.59 years | Mean: 8.4 years<br>SD: 7.1 years | Mean: 8.4 years<br>SD: 7.99 years | Mean: 157.6 months<br>SD: 201.6 months<br>Median: 87 months<br>Range: 5-535 months | NA |
| **Baseline SELENA-SLEDAI** | Median: 10.5<br>Range: 8-21 | Median: 12<br>Range: 8-27 | Median: 11<br>Range: 8-22 | Median: 10<br>Range: 6-16 | Mean: 10.8<br>SD: 3.9 | Mean: 11.3<br>SD: 3.37 | Mean: 10.3<br>SD: 3.2 | Mean: 10.4<br>SD: 3.26 | Mean: 1.0<br>SD: 1.7<br>(uses SLEDAI, not SELENA-SLEDAI) | Median: 6<br>Range: 0-22<br>Mean: 7.5 |

Abbreviations: ITT, intention to treat; MIU, million international units; NA, not available; PP, per protocol; RCT, randomized-controlled clinical trial; SD, standard deviation; SELENA-SLEDAI, Safety of Estrogens in Lupus National Assessment modification of Systemic Lupus Erythematosus Disease Activity Index; SRI-4, Systemic lupus erythematosus responder index 4.

**Table 2:**
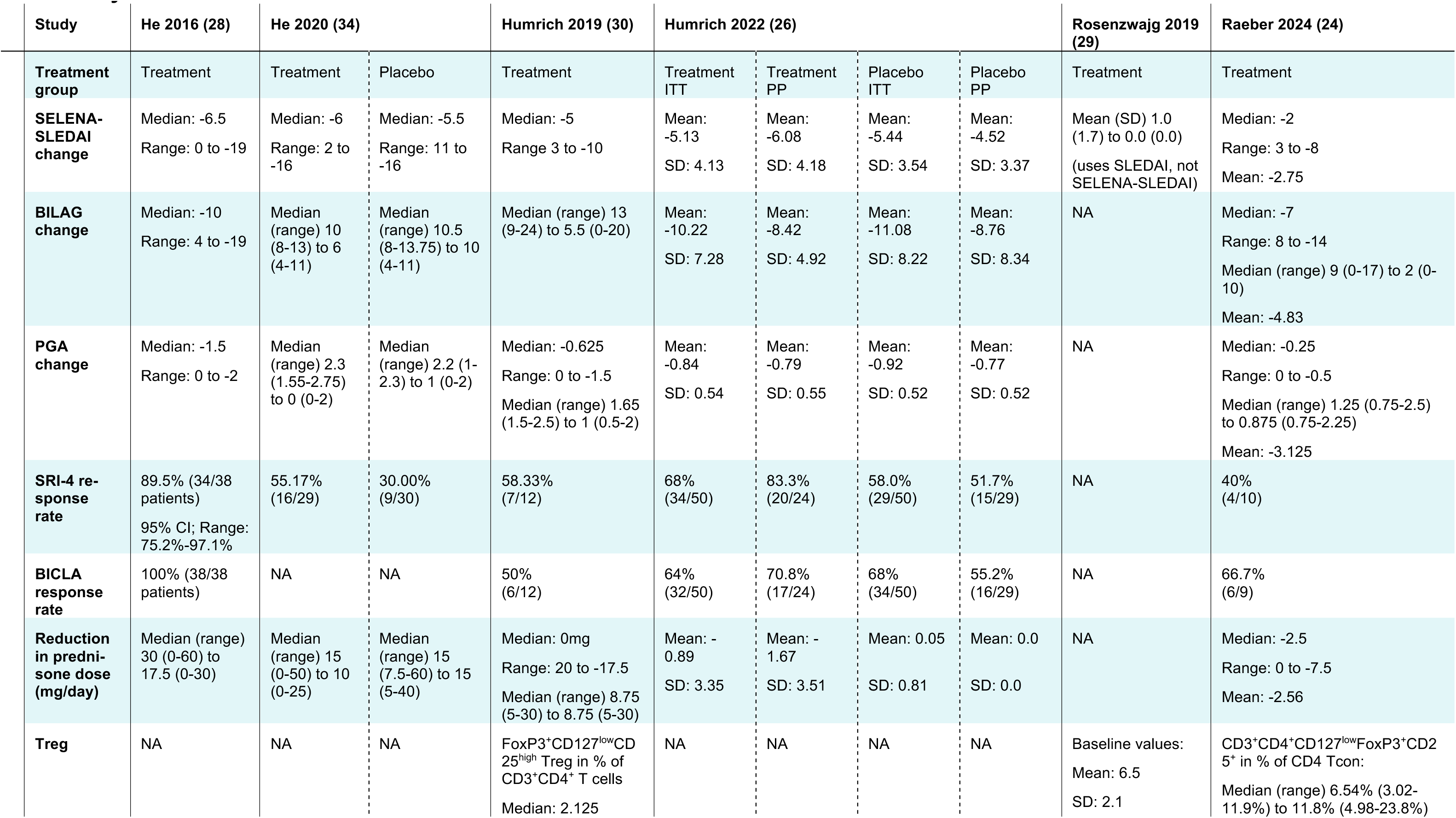
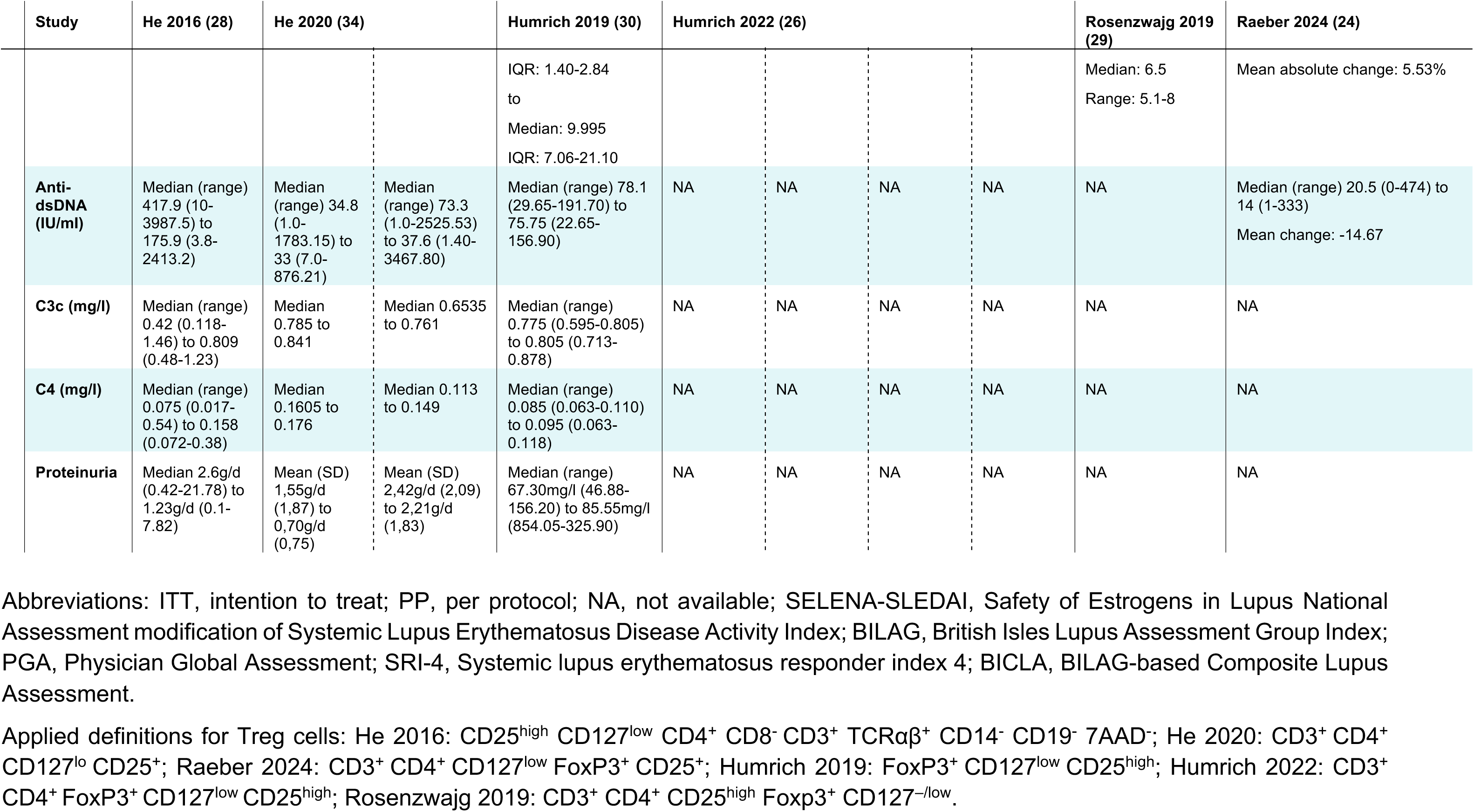
Synthesized results.

**Table 3:**
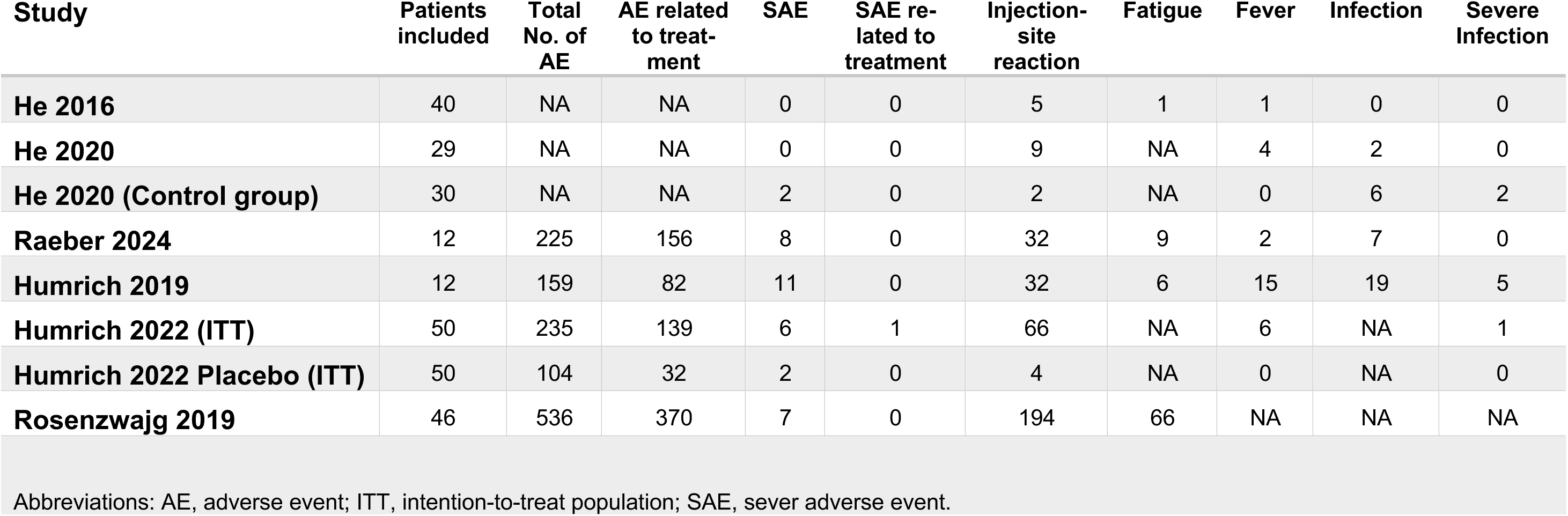
Summary of adverse events.

| **Study** | **Patients included** | **Total No. of AE** | **AE related to treatment** | **SAE** | **SAE related to treatment** | **Injection-site reaction** | **Fatigue** | **Fever** | **Infection** | **Severe Infection** |
| --- | --- | --- | --- | --- | --- | --- | --- | --- | --- | --- |
| **He 2016** | 40 | NA | NA | 0 | 0 | 5 | 1 | 1 | 0 | 0 |
| **He 2020** | 29 | NA | NA | 0 | 0 | 9 | NA | 4 | 2 | 0 |
| **He 2020 (Control group)** | 30 | NA | NA | 2 | 0 | 2 | NA | 0 | 6 | 2 |
| **Raeber 2024** | 12 | 225 | 156 | 8 | 0 | 32 | 9 | 2 | 7 | 0 |
| **Humrich 2019** | 12 | 159 | 82 | 11 | 0 | 32 | 6 | 15 | 19 | 5 |
| **Humrich 2022 (ITT)** | 50 | 235 | 139 | 6 | 1 | 66 | NA | 6 | NA | 1 |
| **Humrich 2022 Placebo (ITT)** | 50 | 104 | 32 | 2 | 0 | 4 | NA | 0 | NA | 0 |
| **Rosenzwajg 2019** | 46 | 536 | 370 | 7 | 0 | 194 | 66 | NA | NA | NA |

Abbreviations: AE, adverse event; ITT, intention-to-treat population; SAE, sever adverse event.

### b. Study characteristics and synthesis of findings

#### Humrich et al. (2019)

Humrich et al.’s (2019) study on the effects of using low-dose IL-2 to treat SLE was an investigator-initiated, single-center, open-label phase 1 and 2a clinical trial with 12 patients (30). Patients received 4 cycles of low-dose IL-2 (Aldesleukin, Novartis Pharma) injected subcutaneously daily on 5 consecutive days; the second cycle proceeded 9 days after the first, whereas 16-day breaks from treatment separated the remaining cycles. The standard dose was 1.5 million international units (MIU) of IL-2 during the first cycle and 3.0 MIU of IL-2 in the following cycles, with the possibility of increased and decreased dosing depending on the expansion of Treg cells and adverse effects (average cumulative dose = 32.6 MIU). The study met its primary endpoints of safety and of a 100% increase in CD25^high^ Treg cells after 4 treatment cycles at Day 62 in 11 of 12 patients (fold-change of 3.72 compared with baseline, p = 0.0005, Wilcoxon signed-rank test). Of 159 adverse events identified according to the Common Terminology Criteria for Adverse Events version 4.03, 75 (47%) were treatment-related, and most were graded mild to moderate (i.e., grades 1 to 2). However, no serious adverse events were recorded during the treatment period. Concerning secondary clinical endpoints comparing the baseline to Day 62, the following median improvements were observed: a drop from 10.0 (interquartile range [IQR] 8.0–13.5) to 5.0 (IQR 4.0–11.0; p = 0.0103, Wilcoxon signed-rank test) in the Safety of Estrogens in Lupus Erythematosus National Assessment version of the SLEDAI (SELENA-SLEDAI) score; shifts in the BILAG-2004, including in category A from 7 to 4 patients, in category B from 11 to 3 patients, in category C from 9 to 11 patients, and in category D from 0 to 7 patients; and a drop from 1.7 (IQR 1.5–2.0) to 1.0 (IQR 0.5–1.9) in PGA scores. Altogether, the trial revealed the significant expansion of Treg cells following treatment with IL-2, which was safe and tolerated well, and suggests the treatment’s potential clinical benefit of reducing disease activity.

#### He et al. (2016)

In He et al.’s (2016) single-center, open-label study testing treatment with low-dose IL-2, 40 patients with SLE and a control group of patients with SLE fulfilling the same inclusion and exclusion criteria were recruited to receive the standard of care (28). Because the control group was recruited 19 months after the completion of the experimental IL-2 treatment, the study is not registered as a controlled trial. Patients received 3 cycles of low-dose IL-2 (recombinant human IL-2^Ser125^, Beijing SL Pharma, China); each cycle consisted of 1 MIU applied every other day for 2 weeks (i.e., 7 injections), followed by a 2-week break from treatment (cumulative dose = 21 MIU). In 27 patients with available blood samples, a significant increase in CD25^high^ Treg cells within the total of CD4^+^ T cells was observed between baseline and Week 12 (p < 0.001, paired *t* test). In 38 evaluable patients, SELENA-SLEDAI scores showed a median reduction by −6.5 (range: 0 to −19) when comparing the baseline to Week 12 (p < 0.001, paired *t* test), which coincided with an SRI-4 response observed in 89.5% of patients at Week 12 (95% confidence interval [CI]: 75.2–97.1%). Of 38 patients evaluated for adverse events, five exhibited injection-site reactions and one showed fatigue and one fever. Overall, with few adverse events reported, the results of the open-label trial suggest the clinical benefits of using low-dose IL-2 to treat SLE.

#### Rosenzwajg et al. (2019)

In Rosenzwajg et al.’s (2019) open-label, multi-center phase 1–2a TRANSREG basket trial testing low-dose IL-2 in 11 autoimmune diseases (32), results for six patients with SLE were reported. Patients received 5 consecutive injections of 1 MIU of IL-2 (Aldesleukin, Novartis) from Day 1 to Day 5, followed by 1 injection of 1 MIU of IL-2 every 2 weeks until Day 180 (cumulative dose = 17 MIU). Treg cells increased by 2.5-fold (standard deviation [SD] 0.7) from baseline to Day 8, although the authors did not provide a statistical analysis for the SLE subgroup. The SLEDAI score decreased from 1.0 (SD 1.7) at baseline to 0.0 (SD 0.0) at Day 183. Across all 46 patients representing 11 autoimmune diseases, 370 non-serious adverse events and zero serious adverse events related to the treatment were reported. The trial’s findings thus support the safety of treatment with low-dose IL-2 as well as the expansion of Treg cells. Even so, conclusions on the treatment’s efficacy for SLE are limited by the small sample size and low disease activity at baseline.

#### Raeber et al. (2024)

The results of our own investigator-initiated, open-label, single-center phase 2 clinical trial have recently been made available (24). Therein, 12 patients with SLE received 4 cycles of 1.5 MIU of IL-2 (Aldesleukin, Novartis) daily for 5 consecutive days, with a treatment break of 2 weeks after each cycle (cumulative dose = 30 MIU). From baseline to the completion of 4 treatment cycles at Day 68, mean percentages of CD25^+^ Treg cells increased from 7.2% (SD 2.69) to 12.7% (SD 5.3; p = 0.0047, paired *t* test). The expansion of Treg cells coincided with a reduction in the mean SELENA-SLEDAI score from 7.5% (SD 5.7) to 4.8% (SD 5.1; p = 0.0032, Wilcoxon signed-rank test), in the mean BILAG-2004 score from 8.9% (SD 5.5) to 4.0% (SD 3.8; p = 0.0148, paired t test), and in the mean PGA score from 1.4 (SD 0.46) to 1.1 (SD 0.46; p = 0.0020, Wilcoxon signed-rank test). Clinical adverse events were generally mild and transient. The results of the trial thus confirm the findings of previous trials showing the expansion of Treg cells by low-dose IL-2, a clinical treatment response, and the overall safety of treatment with low-dose IL-2.

#### He et al. (2020)

In their randomized, double-blind, and placebo-controlled phase 2 trial, He et al. (2020) enrolled 30 patients with SLE to receive low-dose IL-2 and 30 patients to receive a placebo (27). The primary endpoint was a significant difference in SRI-4 response at Week 12. Patients received 1 MIU of IL-2 (recombinant human IL-2^Ser125^, Beijing SL Pharma, China) daily for 2 weeks, followed by a 2-week break from treatment for a total of three treatment cycles (cumulative dose = 21 MIU). The trial missed its primary endpoint, with a reported SRI-4 response of 55.2% in the IL-2 group and 30.0% in the placebo group (p = 0.0520, paired *t* test). Improvements in the SELENA-SLEDAI, BILAG-2004, and PGA scores were reported and are shown in Table 2. Flu-like symptoms, fever, and injection-site reactions were more commonly reported in the IL-2 group than the placebo group, although no serious adverse events were reported in either group. Despite missing its primary endpoint, the trial showed a trend toward improved clinical response in IL-2-treated patients with SLE compared with placebo.

#### Humrich et al. (2022)

In another randomized, double-blind, placebo-controlled phase 2 trial, Humrich et al. (2022) enrolled 100 patients who were 1:1 randomized to receive low-dose IL-2 versus placebo (26). Patients received subcutaneous injections of either 1.5 MIU of IL-2 (ILT-101, Aldesleukin, ILTOO Pharma) or a placebo daily for 5 consecutive days, followed by a weekly injection from Day 8 to Week 12 (cumulative dose = 18.5 MIU). The primary endpoint of improved SRI-4 response at Week 12 was not met, with a 68% response rate in the IL-2 group and a 58% response rate in the placebo group (p = 0.3439). Due to the surprisingly high response rates (i.e., 100%) in the placebo groups recruited at two study sites in Bulgaria, the authors conducted an additional per-protocol analysis excluding these two sites that revealed a significant difference in the SRI-4 response at Week 12, with an 83.3% response rate in the IL-2 group and a 51.7% response rate in the placebo group (p = 0.0168). Additional clinical results, including SELENA-SLEDAI, BILAG-2004, and PGA scores, are provided in Table 2. Concerning safety, treatment with IL-2 was generally tolerated well, with treatment-related adverse events being mostly transient and of mild to moderate severity.

### c. Data analysis

Altogether, the trials reveal a heterogeneous picture, with SRI-4 responses ranging from 50.0% to 89.5% (weighted mean = 68.7%) in the IL-2 groups and 30.0 to 58.0% (weighted mean = 47.5%) in the placebo groups, with Humrich et al.’s (2022) per-protocol analysis excluded (Fig. 2a, left panel). Due to the relatively few trials recording BICLA responses, the data seem more homogenous in the IL-2 groups, with a range from 50.0% to 77.8% (weighted mean = 64.0%) and a rate of 68% in the placebo group, with Humrich et al.’s (2022) per-protocol analysis excluded (Fig. 2a, right panel). The correlation of changes in SLEDAI and BILAG scores upon treatment with IL-2 according to linear regression did not result in a significant correlation (p = 0.0689), likely due to the small sample size, as only two articles provided sufficient data (Fig. 2b) (24, 30). A comparative analysis of SRI-4 and BICLA responses reveals significant overlap in responses, with a considerable fraction of patients only showing response in one domain (Fig. 2c). Of the 21 evaluable patients in Humrich et al.’s (2019) and Raeber et al.’s (2022) studies, eight patients showed SRI-4 and BICLA responses; however, three patients showed an SRI-4 response only, whereas five patients showed a BICLA response only. Beyond that, five patients showed neither an SRI-4 nor BICLA response (Fig. 2d). Although limited by a small sample size, those findings suggest the higher sensitivity of BICLA scores in detecting clinical response.

**Fig. 2.**
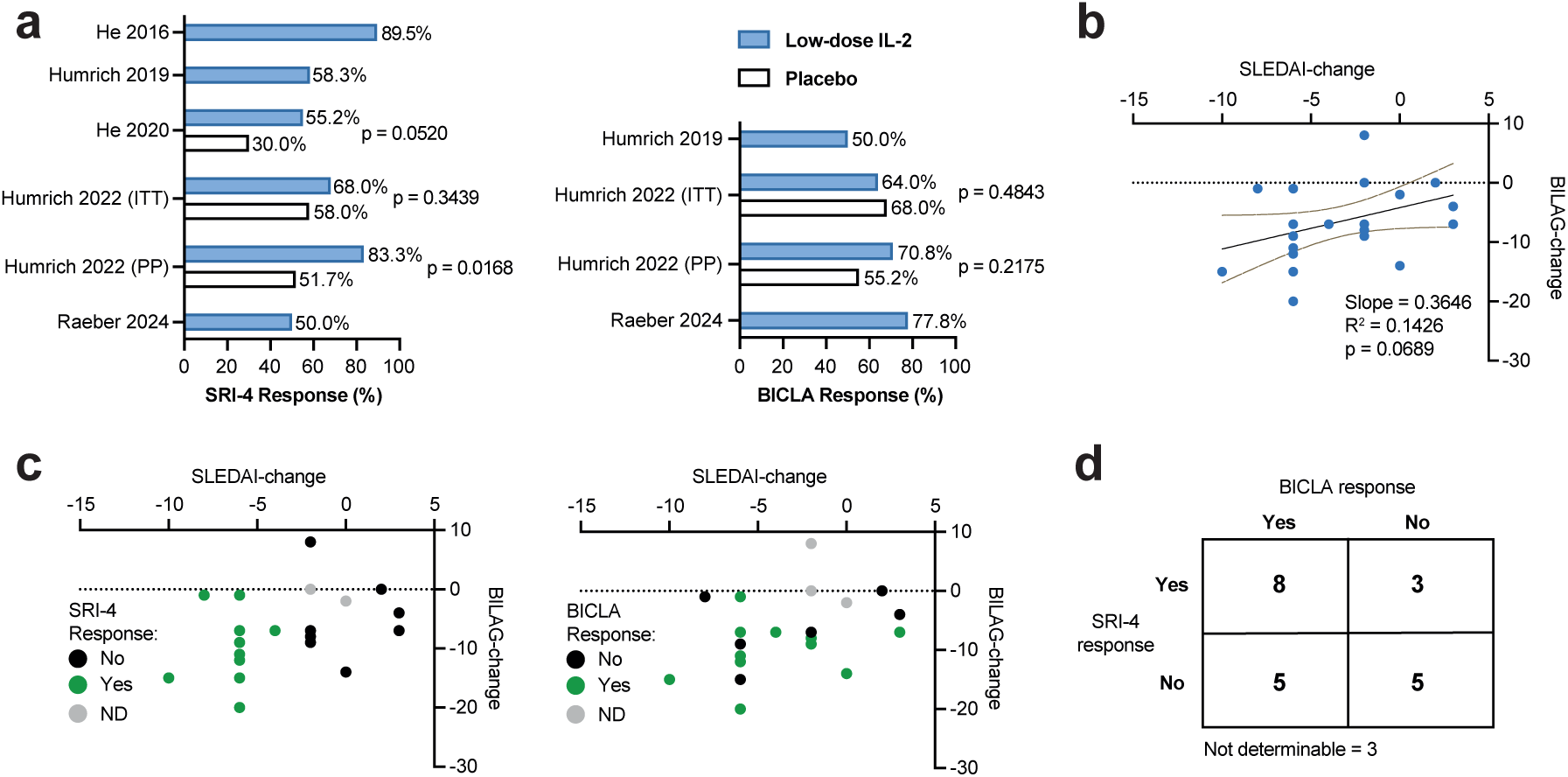
Results from clinical trials testing low-dose IL-2 in SLE. **a**, Extracted SRI-4 and BICLA response. For randomized controlled trials statistical significance of treatment response compared to placebo was extracted from the original publications. **b**, Linear regression with 95% confidence intervals correlating changes in SLEDAI and BILAG upon IL-2 treatment in the Humrich et al. 2019 and Raeber et al. 2024 trials (24, 30). **c and d**, Same data as in b displaying SRI-4 (c, left panel) and BICLA response (c, right panel), and table summarizing patient responses (d).

### d. Risk of bias assessment

Risk of bias was assessed by using the modified Downs and Black checklist shown in Table S2.

## IV. Discussion

### a. Summary of major findings

In six articles, we reviewed four open-label studies and two RCTs testing low-dose IL-2 treatment in patients with SLE. In general, treatment with low-dose IL-2 can be considered to be safe due to only mild to moderate adverse events have been reported (Table 3). None of the trials, which included 150 SLE patients receiving low-dose IL-2, revealed any treatment-related severe adverse event. Concerning response rate, the four open-label trials confirmed the expansion of Treg cells by low-dose IL-2 coinciding with a clinical response. The two RCTs did not reveal significant benefit of treatment with IL-2 versus placebo in patients with SLE. Several factors might have contributed to that failure. For example, the placebo groups in Humrich et al.’s (2022) trial at the two study sites in Bulgaria showed an unusual high response rate of 100%. Furthermore, using a more sensitive response measure such as BICLA instead of SRI-4 might be more suitable to detect responses to IL-2-mediated treatment. Well-designed, larger phase 3 trials are required to determine whether low-dose IL-2 is indeed effective for treating SLE.

### b. Limitations

To our knowledge, our systematic review marks the most comprehensive synthesis of findings about treating SLE with low-dose IL-2 and the first-ever analysis on the sensitivity of different clinical response measures. Even so, our findings have some important limitations. Firstly, the available data represent different outcome measures with partially incomplete data. Secondly, because we included open-label trials, our analysis was prone to bias given the lack of individual control groups.

### c. Conclusions

Despite the recent failure of phase 2 RCTs testing the use of low-dose IL-2 to treat SLE, IL-2 remains an attractive target for the selective expansion of Treg cells. Apart from larger trials that may ultimately show the efficacy of using IL-2 to treat such a heterogeneous, difficult-to-treat disease, improved IL-2 formulations with increased CD25-bias might be even more promising (33).

## V. Data availability statement

The full dataset generated for this study is available upon request from the corresponding author.

## VI. Author contributions

MER: conception of the study. LB and MER: data collection. LB and MER: analysis and interpretation of data. LB, OB, and MER: writing of the manuscript.

## VII. Acknowledgements

This work was supported by the Swiss National Science Foundation (310030-172978 and 310030-200669; to OB), Hochspezialisierte Medizin Schwerpunkt Immunologie (HSM-2-Immunologie; to OB), the Clinical Research Priority Program CYTIMM-Z of University of Zurich (to OB), a Young Talents in Clinical Research Fellowship and a Project Grant by the Swiss Academy of Medical Sciences and G. & J. Bangerter-Rhyner Foundation (YTCR 32/18 and YTCR 08/20; to MER), and a Filling-the-Gap Fellowship by University of Zurich (to MER).

---

**Table S1:**
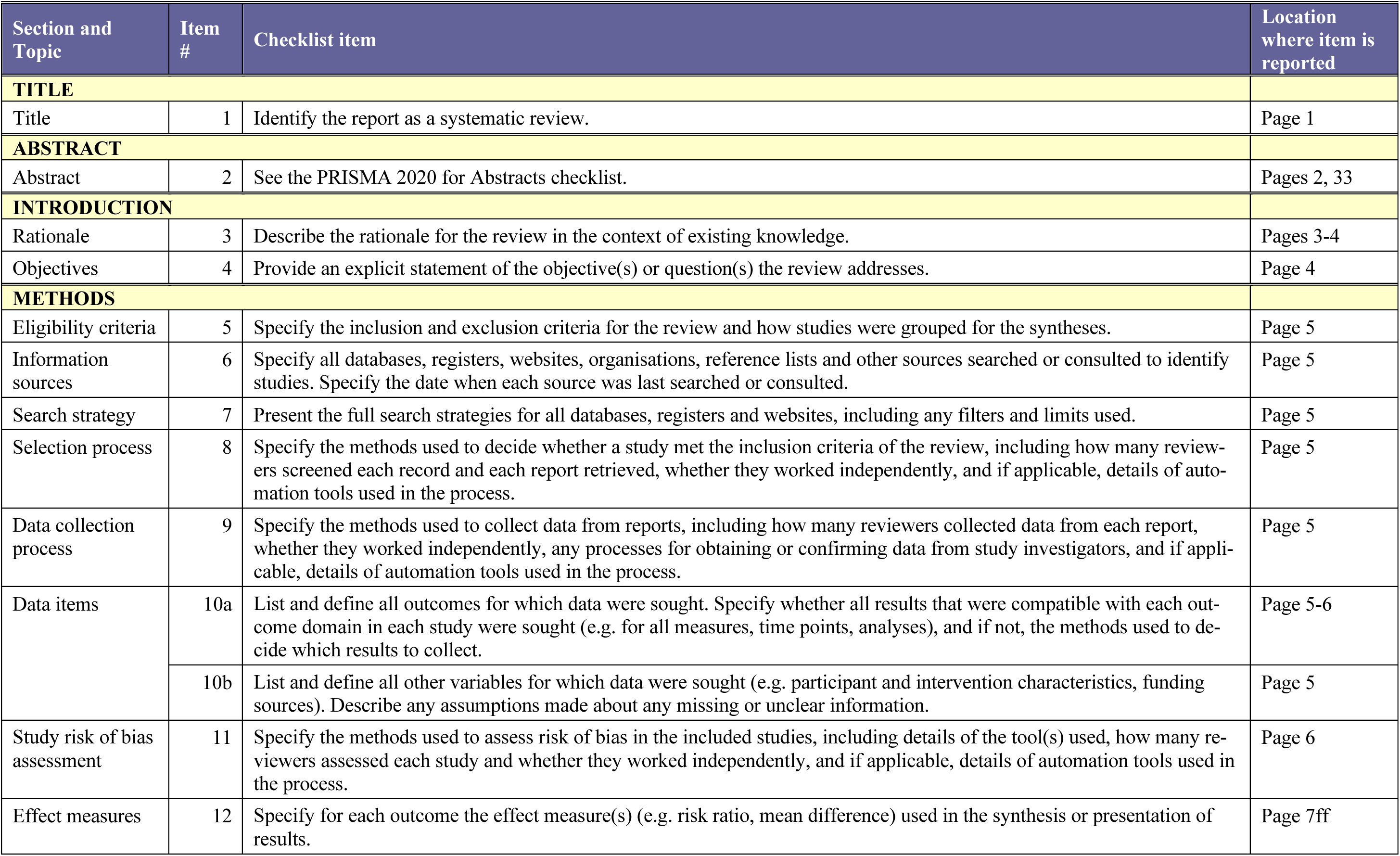
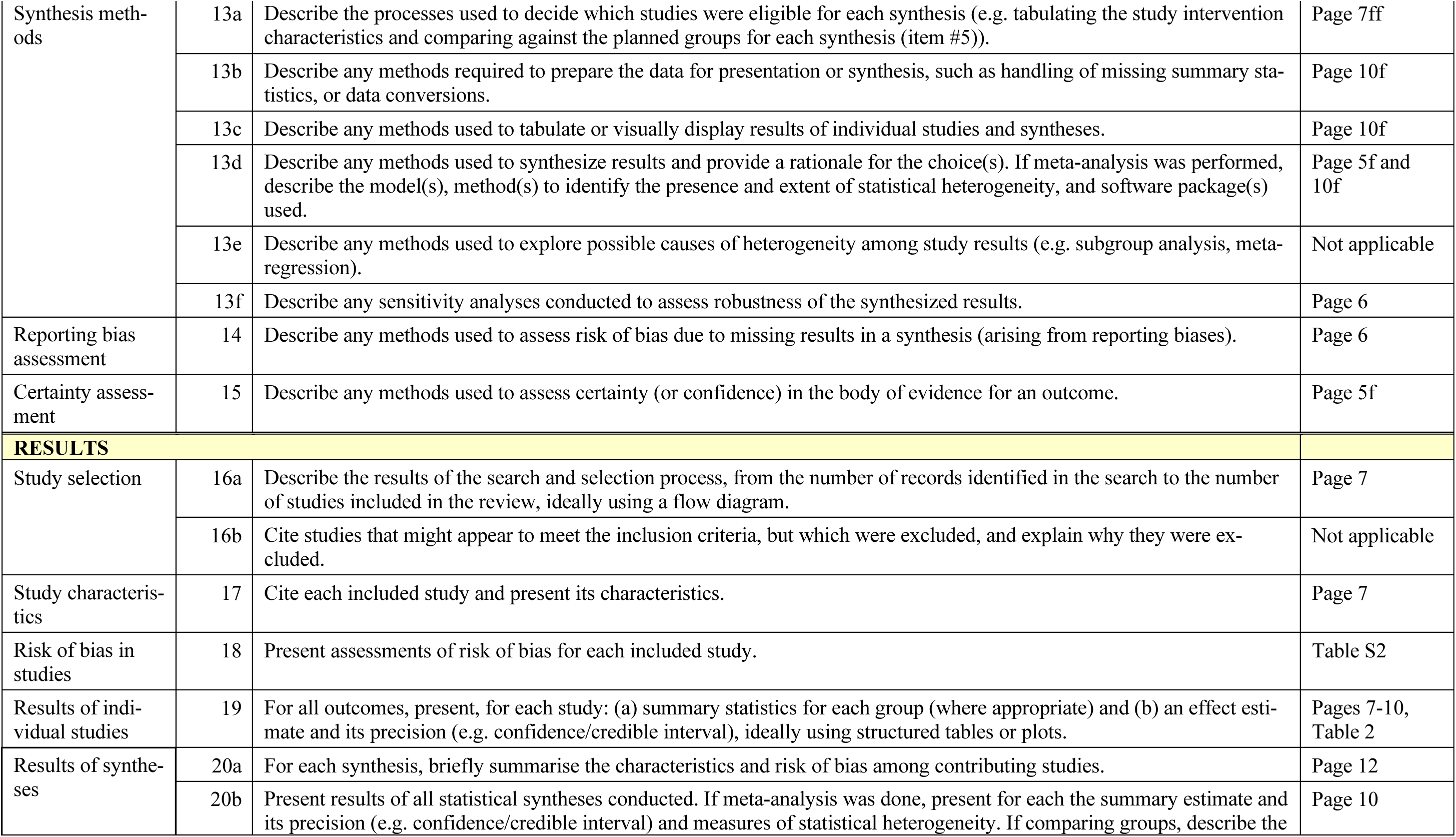
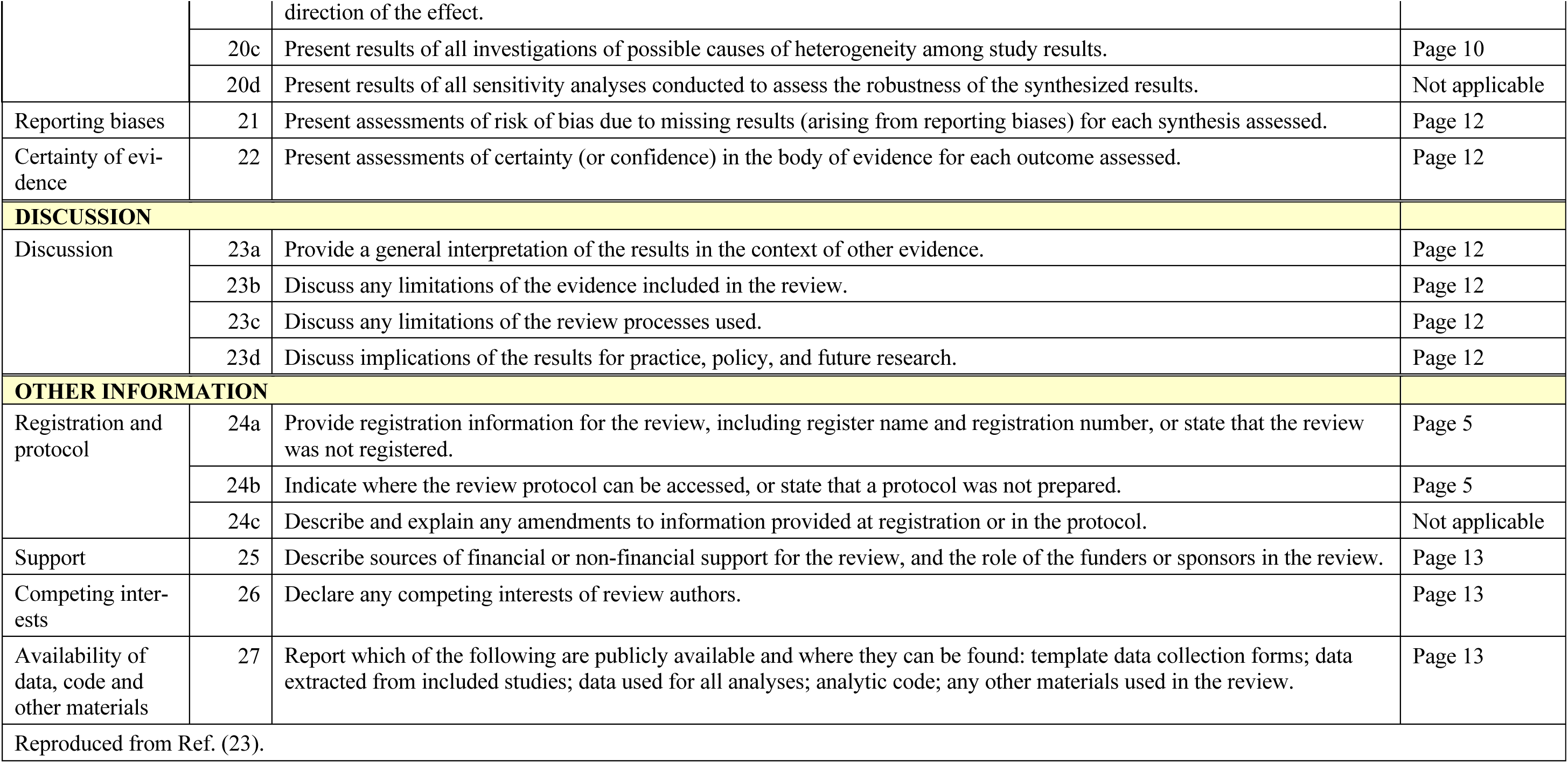
Preferred Reporting Items for Systematic Reviews and Meta-Analyses (PRISMA) checklist.

**Table S2:**
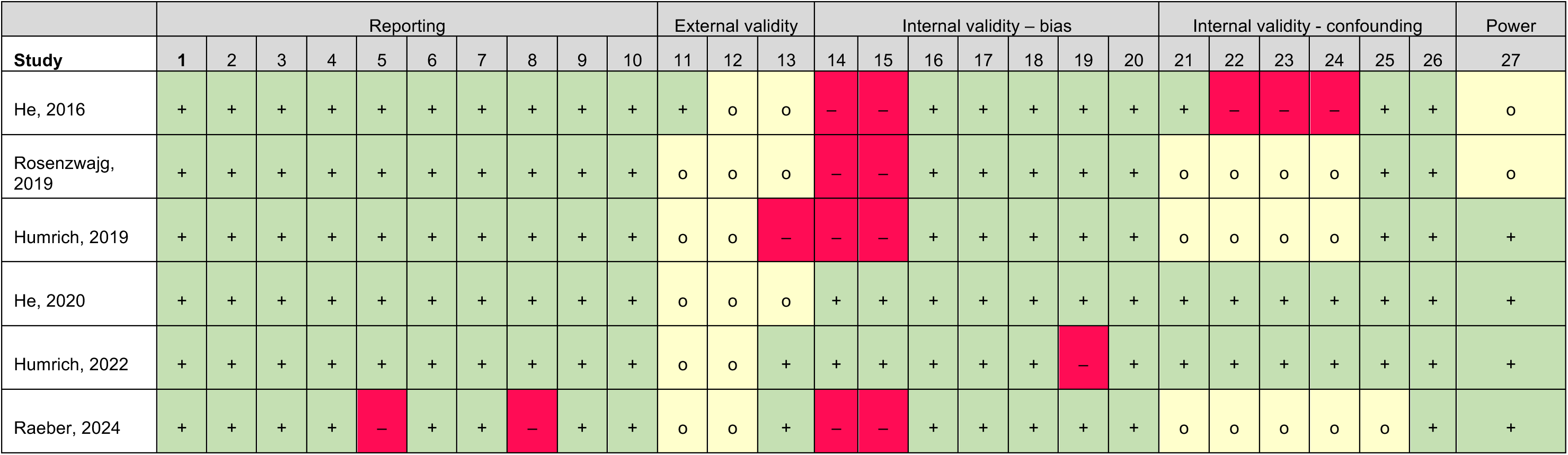
Modified Downs and Black Risk of Bias Assessment.

|  | Reporting |  |  |  |  |  |  |  |  |  | External validity |  |  | Internal validity – bias |  |  |  |  |  |  | Internal validity - confounding |  |  |  |  |  | Power |
| --- | --- | --- | --- | --- | --- | --- | --- | --- | --- | --- | --- | --- | --- | --- | --- | --- | --- | --- | --- | --- | --- | --- | --- | --- | --- | --- | --- |
| Study | 1 | 2 | 3 | 4 | 5 | 6 | 7 | 8 | 9 | 10 | 11 | 12 | 13 | 14 | 15 | 16 | 17 | 18 | 19 | 20 | 21 | 22 | 23 | 24 | 25 | 26 | 27 |
| He, 2016 | + | + | + | + | + | + | + | + | + | + | + | o | o | – | – | + | + | + | + | + | + | – | – | – | + | + | o |
| Rosenzweig, 2019 | + | + | + | + | + | + | + | + | + | + | o | o | o | – | – | + | + | + | + | + | o | o | o | o | + | + | o |
| Humrich, 2019 | + | + | + | + | + | + | + | + | + | + | o | o | – | – | – | + | + | + | + | + | o | o | o | o | + | + | + |
| He, 2020 | + | + | + | + | + | + | + | + | + | + | o | o | o | + | + | + | + | + | + | + | + | + | + | + | + | + | + |
| Humrich, 2022 | + | + | + | + | + | + | + | + | + | + | o | o | + | + | + | + | + | + | – | + | + | + | + | + | + | + | + |
| Raeber, 2024 | + | + | + | + | – | + | + | – | + | + | o | o | + | – | – | + | + | + | + | + | o | o | o | o | o | + | + |

